# Literature Review Interventions to Anticipate Catheter-Related Urinary Tract Contaminations in Seriously Care Units: Philosophy of Nursing

**DOI:** 10.1101/2023.12.11.23299804

**Authors:** Sri Rusmini, Moses Glorino Rumambo Pandin

## Abstract

**Background:** Urinary tract infections are among the adverse outcomes of urinary catheter placement. After the installation of a catheter, urinary tract infections happen more often than twice a day. Examining prevention strategies for UTIs in critical care units is the aim of this writing audit.

**Method:** Use ProQuest, Scopus, and ScienceDirect to look up review literature. Urinary tract infections, prevention and management, and intensive care unit were the search terms used to find publications. Search parameters for full-text, open-access, English-language papers published between 2019 and 2023.

**Results:** The study’s findings indicate that adherence to patient information recording, installation and maintenance procedures, guidelines for the detection and prevention of CAUTI, staff education and good nursing practices, and the application of CAUTI prevention can all help lower the incidence of urinary tract infections.

**Conclusion:** Healthcare facilities need to have a program in place to avoid urinary tract infections brought on by catheter installation, as well as the best care possible for patients who use them. In order to solve the problem, all interdisciplinary teams must collaborate and analyze the issue using the PDSA methodology.

## Introduction

Urinary catheters are used in critically ill patients to monitor their fluid balance. According to Leon et al (Leone et al., 2004).CAUTI may have a significant role in the increased severity, mortality, expenses, and length of stay in the clinic. Cystitis is a common sign of urinary tract infections, which are often caused by Staphylococcus and E. coli in the urine sample (Birgitta Hovelius, 1984). Urinary tract infection symptoms are easily identifiable and typically include fever (F.M.E. Wagenlehner, E. Loibl, H. Vogel, 2006).

A urinary tract disease is an impairment that affects the kidneys, ureters, bladder, and urethra, as well as other components of the urinary system. 75% of nosocomial infections are typically associated with urinary catheter use. Urinary catheters can be inserted in as many as 15–25% of hospitalized patients during their stay. Long-term use of a urinary catheter is one risk factor associated with catheter configuration. According to the Centers for Disease Control and Prevention (2015), it should be used as if it were coordinated and eliminated when not needed.

By reducing unnecessary catheter use, shortening catheter use times, and advancing addition techniques, urinary tract disorders can be predicted. Urinary tract infections can be reduced by healthcare professionals, and interventions that target their behaviors are essential for optimal understanding and treatment (Wanat et al., 2020). The evaluation, mediation, prediction, and control rate of urinary tract disorders in critically ill units is the motivation behind this writing survey.

## Method

In order to predict and manage urinary tract infections in critical care units, this study used a writing audit approach. His publication period from 2019 to 2023 is one of the publication requirements. This article makes use of full text and original research. When searching the literature with ProQuest, ScienceDirect, and Scopus, The terms “urine tract infections,” “prevention and control,” and “intensive care units” were used in the literature search. After looking through the 736 publications that the keyword search produced, we looked through the inclusion criteria and found 6 research papers.

**Figure 1.**
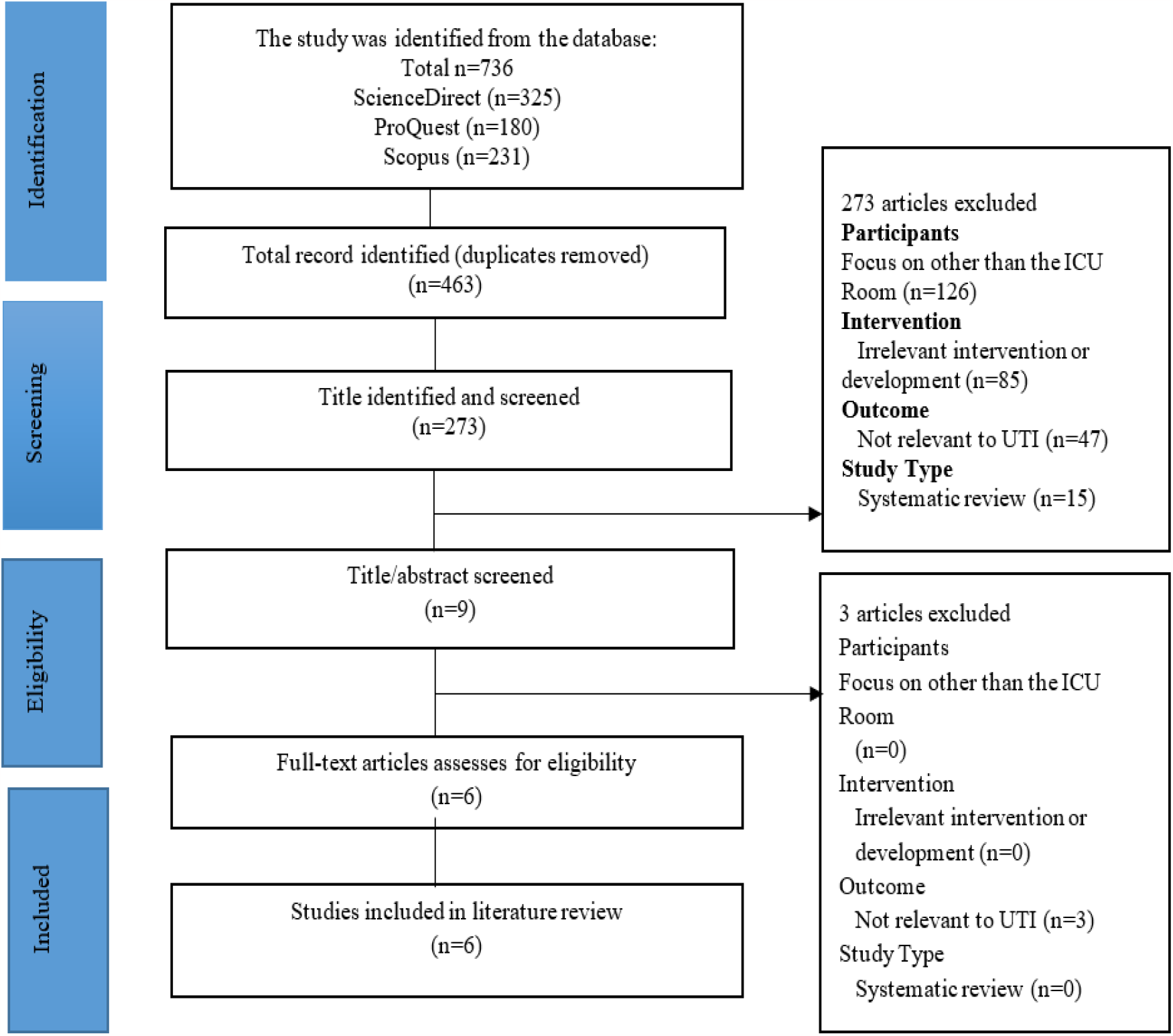
Flowchart for PRISMA.

**Tabel 1.**
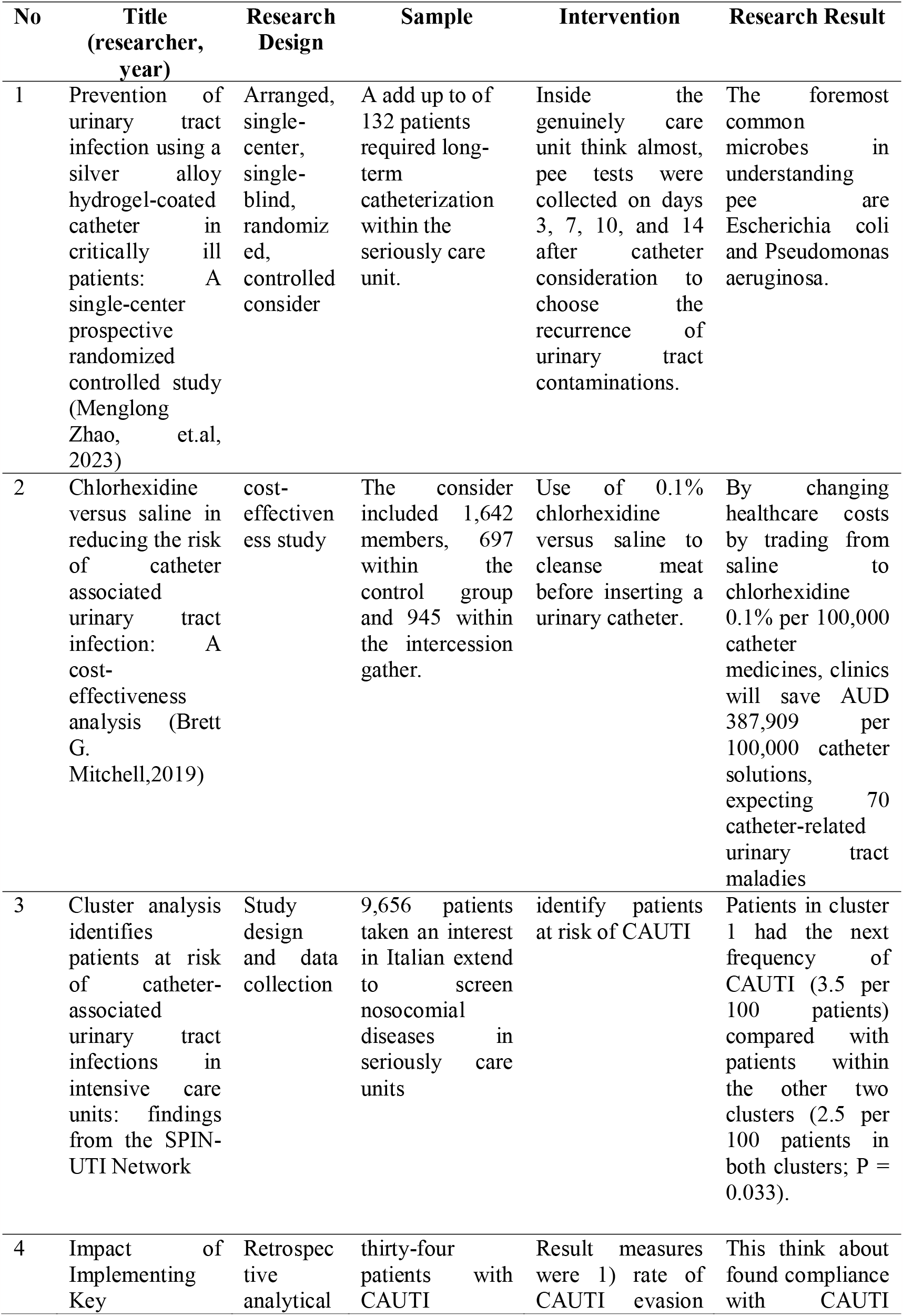
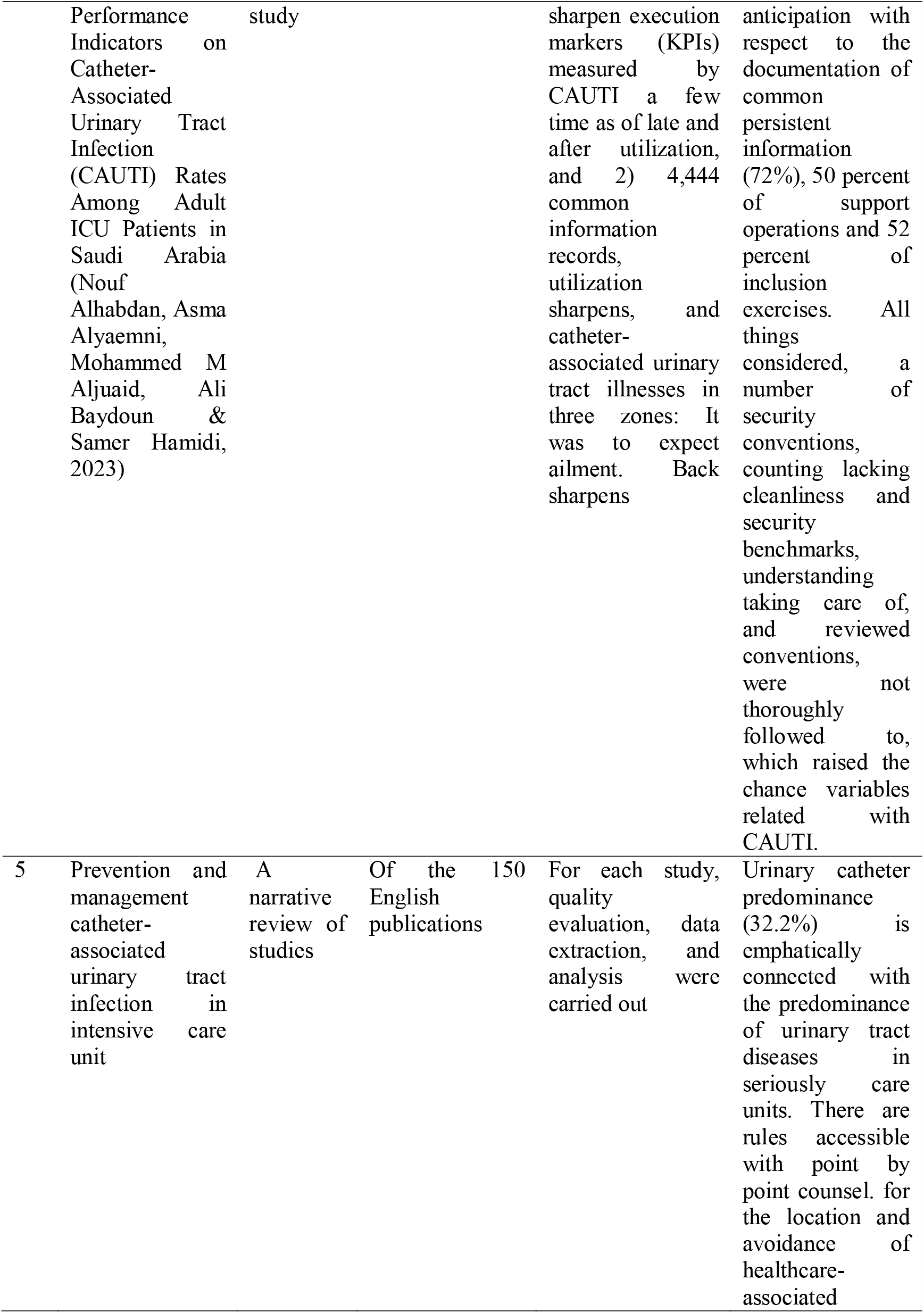
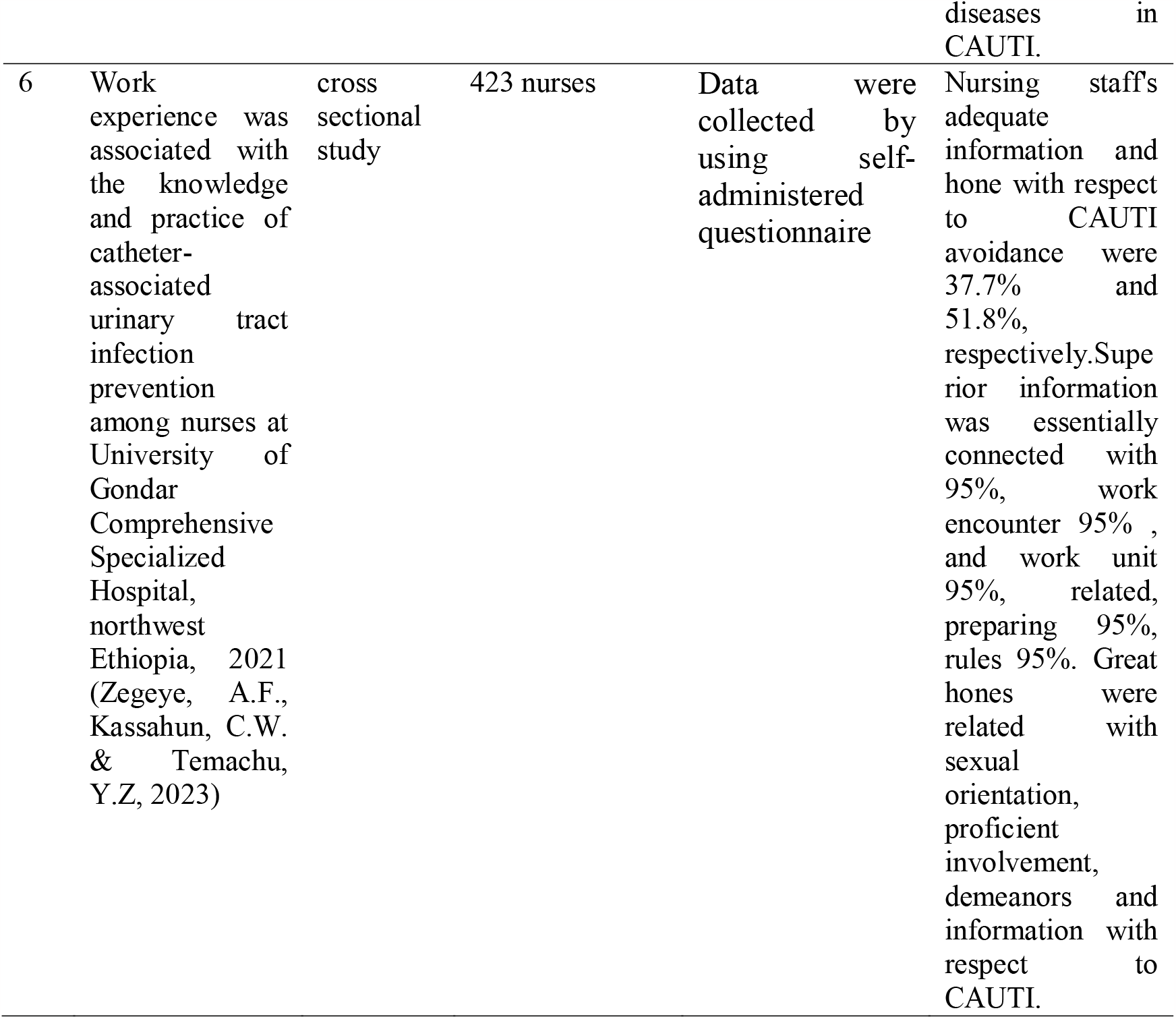
Search results for articles on preventing urinary tract infections in the ICU.

## Results and Discussion

### 1. Catheter-Associated Urinary Tract Contamination Avoidance Program

Programs to prevent urinary tract infections include limited use of urinary catheters, placing catheters in sterile environments, and promoting a closed seepage framework (Laurie J. Conway, 2012). The most effective prevention technique is to avoid unnecessary catheter insertion and remove the catheter as soon as it is practical (Tenke et al., 2017). According to Fink et al. (Fink et al., 2012), precautions include wearing gloves, cleaning your hands frequently, keeping your PPE sterile, providing daily urinary tract care, teaching new caregivers aseptic procedures and precautions, documenting everything you do, and conducting routine monitoring.

In order to prevent and control urinary tract infections, it is important to use the right catheters, have them installed by trained personnel using aseptic techniques and sterile equipment, and maintain good urinary catheters with a closed, sterile drainage system to prevent obstructions in urine flow (Meddings et al., 2014). It is advised that patients receive care using aseptic technique as a standard. In order to adopt suitable risk mitigation strategies, prevention programs and risk factors need to be assessed. According to Ling et al. (Ling et al., 2023), risks considered included the placement of urinary catheters without necessity, placing them for an extended period of time, maintaining aseptic technique during catheter placement and maintenance, failing closed drainage systems, obstructing urine flow, and placing catheters poorly. The following are suggestions for preventing UTIs: (1) Provides personnel with knowledge of the etiology and risk factors for infection.

Programs to prevent CAUTIs are designed to reduce unnecessary catheter setup and shorten catheter usage times. Additionally, it’s made to make sure you adhere to his CAUTI prevention program, which consists of the following: (1) staff available for training in insertion and withdrawal techniques and treatment procedures; (2) highly qualified and experienced personnel present; (3) sterile equipment available for catheter installation and maintenance; (4) CAUTI screening programs set up to reduce unnecessary catheter use and minimize catheter course of action; (5) instructions for catheter placement; (6) personnel trained in use and result monitoring; and (7) monitoring implementation (Ling et al., 2023). Suggestions for the creation of CAUTI prevention initiatives.

### 2. Surveilans

One way to prevent infections is to keep an eye on urinary tract infections linked to health issues (Bagchi et al., 2020). Longer catheterizations, hospital stays, and ICU stays put patients at risk for infection and necessitate closer observation (Talaat et al., 2010). Regulations and risk-based assessments are used in healthcare settings to monitor urinary tract infections. CAUTI Monitoring Suggestions: (1) employing conventional techniques; (2) it is not advised to routinely screen catheter-using individuals for the occurrence of asymptomatic bacteriuria. (3) Give clinical nursing personnel and nursing staff feedback when you are monitoring. (4) Notify the treating and supervising physicians of the monitoring outcomes (Ling et al., 2023).

### 3. Carry out control of catheter-related urinary tract contamination

The success of preventing urinary tract infections in healthcare is a result of improvements in the quality of care that lessen the risk of infection. If all interdisciplinary teams collaborate and employ the PDSA technique to identify and address issues, programs can be implemented successfully (Ling et al., 2023). Strong clinical implementation and assessment procedures that prioritize patient safety lead to much better patient experiences and health outcomes. The basis for ongoing progress is the provision of data on urinary tract infections (CAUTI) (Parker et al., 2017). Patient safety can be enhanced by clinical practice through implementation and evaluation. It is anticipated that this will have a major positive impact on patient outcomes and healthcare experiences. Establishing baseline data will serve as a basis for guaranteeing ongoing enhancement and standardizing best practices.

## Conclusion

Review of the literature with a focus on intervention trials that try to lower UTIs caused by urinary catheter implantation. This study can demonstrate how better knowledge of monitoring and infection prevention and control initiatives will enhance patient safety.

## Data Availability

All data produced in the present work are contained in the manuscript

